# Neural stress processing, glucocorticoid functioning, and body mass in lean to obese persons with multiple sclerosis

**DOI:** 10.1101/2022.12.28.22284002

**Authors:** Lil Meyer-Arndt, Jelena Brasanac, Stefanie Gamradt, Judith Bellmann-Strobl, Lukas Maurer, Knut Mai, Joachim Spranger, Tanja Schmitz-Hübsch, Friedemann Paul, Stefan M. Gold, Martin Weygandt

## Abstract

**Background and Objectives:** Obesity aggravates disease severity in multiple sclerosis (MS). Altered neural processing of food motivation and cognitive control, and the sensitivity of these processes to stress have been recognized as key obesity mechanisms but never been investigated in MS.

**Methods:** In this cross-sectional observational study, we evaluated the link between body mass and neural, endocrine and immunological stress parameters in persons with MS (PwMS). We conducted an Arterial-Spin-Labeling MRI task comprising a rest and stress stage (mental arithmetic plus evaluative feedback) in 57 PwMS (37 female, 46.4 ± 10.6 years) covering the full spectrum of the Body Mass Index (BMI [kg/m^2^]; 6 obese, 19 over-, 28 normal-, 4 underweight). We tested whether BMI in MS links to (i) functional connectivity (FC) between stress-reactive brain regions (showing activity differences for stress vs. rest) computed separately for the task’s rest and stress stage, (ii) T cell glucocorticoid sensitivity and (iii) salivary cortisol secretion.

**Results:** BMI correlated positively with MS relapse rate (t = 3.23, p = 0.003 = p_Family-Wise-Error [FWE]-corrected_ = 0.012, and f^2^ = 0.22) and rest stage FC between right anterior insula and supramarginal gyrus (t = 4.02, p = 2.5 · 10^−4^ = p_FWE_ = 0.034, f^2^ = 0.51) and negatively with stress stage FC between right superior parietal lobule and cerebellum exterior (t = -3.67, p = 3.3 · 10^−4^ = p_FWE_ = 0.045, f^2^ = 0.30). Further, BMI was negatively associated with the expression of the co-chaperone FKBP4 on CD8^+^ T cells (t = -2.96, p = 0.003 = p_FWE_ = 0.024, f^2^ = 0.13) and positively with that of FKBP5 (t = 1.83, p = 0.003 = p_FWE_ = 0.024, f^2^ = 0.38).

**Conclusion:** Our study shows that higher BMI in MS is linked to increased FC between key food motivation and stimulus salience regions and to reduced FC between regions critically involved in cognitive control and generation of stressful states. We further report on correlations between BMI and co-chaperones modulating immune system stress responsivity. Taken together, these results demonstrate that BMI in MS is tied to stress processing across different biological systems.

## Introduction

Obesity affects the disease course of multiple sclerosis (MS) in various ways. For example, comorbid obesity in MS is accompanied by elevated disability, central nervous inflammation (1), and relapse rate (2) and clinically isolated syndrome is more likely to convert to MS in obese persons (2). Further, obesity in childhood and adolescence, especially in women, is a risk factor for developing MS (3).

MS obesity research has so far focused on pathomechanisms potentially overlapping in both diseases such as an increased secretion of proinflammatory cytokines by fat cells or of immunomodulatory adipokines including leptin, which upregulate proinflammatory cytokines, or a shift towards proinflammatory T helper cells in obesity (4). Neurobehavioral factors considered as highly relevant obesity mechanisms in endocrinology (see our recent review [5]) and neuroscience (6) were, however, not investigated. Among these, heightened incentive salience, impaired goal-directed control, and maladaptive stress-processing are particularly important. Incentive salience is a motivational process primarily realized by ventral striatum, anterior cingulate cortex, and insular cortex, which can trigger excessive food search and intake after exposure to context stimuli of food consumption (e.g., a visual food percept) as these stimuli acquire reward-like properties through repeated food coupling (6 - 8). In goal-directed control, prefrontal and parietal areas model action consequences and inhibit choices (e.g., eating Pizza) predicted to yield unfavorable consequences by attenuating the impact of regions favoring immediate reward (6, 9). The goal-directed system can downregulate the incentive salience system (10).

Clinical studies emphasize the importance of maladaptive stress processing by showing that obese persons (i) with higher stress tolerance maintain weight loss longer (11), (ii) have a higher risk to increase weight under stressful conditions (12), and (iii) have elevated cortisol levels (13). Simultaneously, an association between a glucocorticoid (GC) receptor gene marker and abdominal obesity (14), and an altered expression of FK506-binding protein 5 (FKBP5; a co-chaperone modulating the intracellular activity of the GC receptor [15]) after dexamethasone application in overweight persons (16) indicates that GC sensitivity contributes to obesity. Mechanisms by which biological stress processing can contribute to obesity are manifold. Corticotropin releasing hormone (CRH) secreted by the hypothalamic-pituitary adrenal axis rapidly suppresses food intake after stress onset whereas CRH-triggered cortisol release stimulates it later (16). Ultimately, one can learn that palatable food intake reduces stress via reinforcement learning and thus establish stress-induced eating as maladaptive coping behavior (17). This is a mechanism primarily mediated by GC and CRH receptors in hippocampus, amygdala, insula, anterior cingulate and prefrontal cortex (18). Moreover, several findings link stress to incentive salience and goal-directed control. Dallmann (18) reports that cortisol intensifies incentive salience by stimulating amygdala neurons projecting to the ventral striatum and Jastreboff and colleagues (19) show that context stimuli of stress and food elicit activity in overlapping striatal, insular, and hypothalamic regions in obese persons. Finally, following (20), stress reduces the inhibitory effect of brain regions promoting self-control/controlled food-choices on regions favoring immediate reward within the goal-directed system by weakening these regions’ functional connectivity (FC).

We here investigated associations between Body Mass Index (BMI; kg/m^2^) and (i) MS severity, (ii) neural stress processing, and (iii) GC functioning in persons with MS (PwMS). First, we evaluated whether BMI is linked to relapse rate, clinical disability, whole-brain grey matter (GM) fraction and T2-weighted lesion load in 57 PwMS. Following (2), we hypothesized that BMI correlates positively with relapse rate. Second, we evaluated associations between BMI and neural stress processing by testing whether BMI is related to rest and stress stage FC of brain regions showing a significant stress vs. rest activity difference (i.e., stress-sensitive regions) using an Arterial-Spin-Labeling (ASL) functional MRI (fMRI) mental math stress task. FC was chosen as neural processing marker due to the pivotal role of regional interplay for obesity (e.g., 5, 9, 20, 21). ASL was selected as it is a quantitative MRI method measuring regional cerebral blood flow (rCBF; ml/100g/min) which is increasingly used in neuroscientific FC studies due to its higher robustness to slow fMRI signal noise relative to that of the alternative technique blood-oxygenation-level-dependent (BOLD) fMRI (22). The task has already been used in a variety of stress studies (22 – 27). Please see also the review (28) and our Discussion on using ASL for studying FC in clinical and cognitive neuroscience. We hypothesized that FC during stress is related to BMI. Third, we tested possible links between BMI and (i) diurnal cortisol secretion and (ii) T cell GC sensitivity in patient sub-samples (cortisol N = 34; GC sensitivity N = 37) and hypothesized that BMI is related to both.

## Materials and methods

### Participants

The study combines data from two study projects carried out by the Experimental and Clinical Research Center and the NeuroCure Clinical Research Center at Charité - Universitätsmedizin Berlin, which included a clinical and an MRI visit at most two weeks apart. Stress fMRI data from the first project were previously published by (24 – 26), those of the second in (23). Comparable inclusion criteria were applied across projects: 2010 McDonald Criteria (29) for relapsing-remitting or secondary-progressive MS in the first project and criteria for relapsing-remitting in the second; continuous disease-modifying treatment (DMT) for at least six (first project) or three (second project) months prior to study participation or no DMT in this period; age 18 years or older; ability to operate the task devices in an unrestricted fashion. Exclusion criteria comprised MRI contraindications, an additional neurologic disorder or acute MS relapse, steroid treatment in the four weeks prior to participation, and an insufficient anatomical image quality assessed visually by M.W. in both projects. Additionally, participants were excluded in case of a known mental health diagnosis (first project) or if a psychotherapist identified a mental health illness other than a depressive or anxiety disorder (second project). Further, we excluded the data of those 16 patients measured in the first project who took part in both projects to guarantee that only one data set was included per individual. Data from the first project were excluded as this was the project with slightly lower anatomical image resolution. From the pooled sample of 60 patients (20 first, 40 second project), we finally removed the data of three patients with pronounced head motion during functional scans indicated by the Framewise Displacement metric (30) as further quality assurance step (see Supplement for details). Consequently, we analyzed data of 57 PwMS in this study.

### Standard protocol approvals, registrations, and patient consents

The projects were conducted in accordance with the Helsinki Declaration of 1975 and approved by the ethics committee of Charité – Universitätsmedizin Berlin (first project: EA1/182/10, amendment V; second: EA1/208/16). Written informed consent was obtained from all study participants.

### Clinical assessments

Neurological impairment was assessed during the clinical visit using the Expanded Disability Status Scale (EDSS; 31). BMI was assessed before brain imaging upon arrival in the imaging center. In particular, the participants’ body weight was measured with a calibrated scale (Kern, Balingen-Frommern, Baden-Württemberg, Germany) and a stadiometer (Seca, Hamburg, Hamburg, Germany) was used for measuring body height. The BDI-II (32) was used in the first project for measuring depressive symptom severity, BDI-I (33) in the second. We merged the results from both measurements into a common metric using the algorithm presented by (34). See (35) for further clinical measures assessed.

### fMRI stress paradigm

The fMRI stress paradigm included seven stages in the first project (1. Rating I, 2. Rest, 3. Rating II, 4. Stress, 5. Rating III, 6. Rest II, 7. Rating IV), while the second project comprised only the first five stages. During the stress stage, the patients had to complete several math tasks. More specifically, in substage 4a (“Evaluation”), we tested the participants’ mental arithmetic performance level and provided feedback in substage 4b (“Feedback”). A nine-point Likert scale with responses ranging from “not at all” to “highly” was used to measure perceived stress during the three rating stages (2 min each). Stage two (8 min) and four (12 min) were used to collect ASL and heart rate data (Supplement). In the present study, we used the data from the first five stages, which were available for both projects. Fig. 1 provides details.

**Figure 1.**
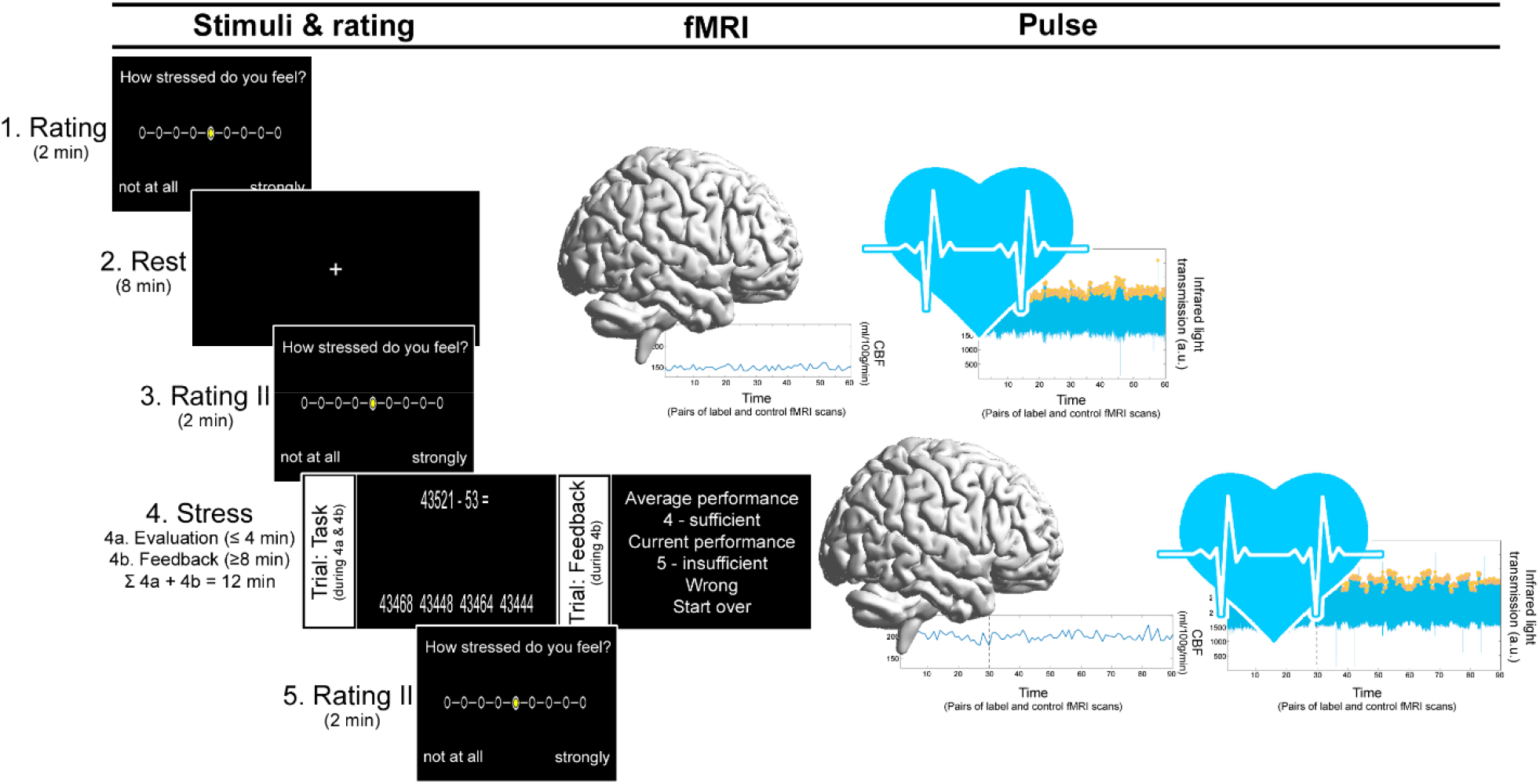
fMRI stress paradigm. The paradigm was divided into five stages. There were two parts to the stress stage: 4a (“Evaluation”) and 4b (“Feedback”). Participants completed subtraction tasks across a number of trials in the Evaluation substage (4a). During each task, patients had to choose the correct solution for equations of the form “operand X minus operand Y” from a list of four possible solutions displayed on a screen with an MRI-compatible response box as quickly as they could. The start value of X was 43521 across all participants. In each of the trials in 4a (and 4b), operand Y was chosen at random (range: 1 – 99). In the first study project, the duration of 4a was 4 minutes or shorter if a participant solved ten tasks correctly earlier. 4a was allocated a fixed duration of 4 min in the second study project. To account for this difference, we included the duration of 4a as CNI “time-to-feedback” in all regression analyses modeling stress fMRI data and evaluated only the rCBF data (and heart rate, cognitive task load or mental arithmetic performance data respectively) collected in the last 8 minutes of stage 4b. The time allotted in 4a to complete a task was 8 seconds. When the participant selected the correct response for a task, X was set to this correct response score in the following task. If the response was inaccurate or too slow, X remained the same. 4a was followed immediately by 4b. 4b was different from 4a in three ways: First, each task included feedback (i.e., German school grades ranging from “1 - very good” to “5 - insufficient”). The difference between the participant’s fastest accurate response in 4a and the response time of a specific trial in 4b was used to select the feedback or school grade respectively. Feedback for incorrect or tardy responses was always “5 - insufficient.” In this instance, X was reset to 43521. Thirdly, according to the participant’s performance, the time for response selection was adjusted. Beginning with at 8 seconds at the start of 4b, this time was decreased by 10% for correct responses and increased by 10% for incorrect or very sluggish answers. This flexible procedure was used to make sure that every participant continuously performed at their highest level possible and that performance was therefore comparable to other participants in terms of an individual performance norm.

### MRI acquisition

Acquisition of MRI scans took place between 3 and 7 pm to control for effects of circadian variations. All MR images were acquired with the same 3 Tesla whole-body tomograph (Magnetom Trio, Siemens, Erlangen, Germany) and standard 12-channel head coil. A pseudo-continuous ASL sequence (22) was used to acquire fMRI scans during rest and stress stage. Field maps were measured for distortion correction of ASL scans. Anatomical images were acquired with T1- and T2-weighted sequences (see Supplement for details).

### MRI preprocessing

#### Anatomical scans

Manual lesion mapping was carried out based on T2-weighted images. The standard preprocessing conducted with SPM12 (Wellcome Trust Centre for Neuroimaging, Institute of Neurology, University College London, London, UK, http://www.fil.ion.ucl.ac.uk/spm) included a spatial mapping of participants’ T2- to T1-weighted scans and a combined spatial normalization and segmentation of T1-weighted scans. During normalization, the T1-weighted images were coregistered to the Montreal Neurological Institute (MNI; 36) standard space. Moreover, we performed computation of tissue-specific group masks and of the whole brain GM fraction based on tissue voxel maps resulting from the segmentation (also see Supplement).

#### Functional scans

Preprocessing of ASL scans was performed with the ACID (37) and ASLtbx (38) toolboxes for SPM12. It included head motion correction, B0 distortion correction, coregistration to the raw T1-weighted images and, subsequently, coregistration to the MNI standard space (voxel resolution 3 · 3 · 3 mm^3^). Mapping to MNI-space was performed by applying the coregistration parameters computed for the T1-weighted anatomical scans described in the section above. Finally, the ASL scans were smoothed in the spatial domain using a three-dimensional gaussian kernel (8 mm full width at half maximum). Afterwards, we extracted rCBF timeseries voxel-by-voxel for all participants and both conditions separately. Data from the (final) eight minutes of both the rest and stress stage were used for all subsequent procedures. We concentrated on the last eight minutes of the stress condition as this time frame was consistently included in the “Feedback” substage 4b across all participants.

### Regional neural stress reactivity and functional connectivity in patients

To provide the source data for the second analysis, we computed parameters of regional neural stress reactivity (defined as stress vs. rest activity differences of regions included in the Neuromorphometrics brain atlas; http://Neuromorphometrics.com) and parameters of pair-of-region-wise FC within patients. Specifically, for testing regional neural stress reactivity, we first generated an rCBF timeseries for each individual, condition, and GM region in the atlas. These timeseries were calculated by averaging the activity of all voxels in a single atlas region that had non-zero rCBF and were covered by the GM group mask. Regions were excluded from all fMRI analyses if they did not contain a single voxel in one or more participants meeting these requirements. 119 out of 122 atlas regions were included (see Supplement for further explanation). By contrasting the regions’ averaged stress vs. rest activity timeseries, regional activity differences were calculated as stress reactivity markers with linear regression for each PwMS. The resulting regression coefficients were used to evaluate regional stress reactivity on the group level in the second analysis (see Supplement for further details). Next, we computed the (Fisher Z-transformed) correlation coefficient for the averaged regional rCBF timeseries of all pairs of areas as parameters of FC per patient separately for the rest and stress condition. FC between regions showing significant stress reactivity was used for testing group level BMI-FC relations in the second analysis.

### Diurnal salivary cortisol

In the second study project, patients received salivettes (Sarstedt, Germany) along with instructions for saliva collection at home. They were instructed to collect saliva once upon waking and once after 9 pm on the two days preceding the MRI scan and submit them during the MRI visit. Samples were then centrifuged for 5 minutes at 1000 × g, aliquoted, and stored at -20 °C until analysis. Following the manufacturer’s instructions, the enzyme-linked immunosorbent assay (ELISA; IBL, Germany) was used to determine cortisol levels. Salivary cortisol data from 34 PwMS were available to compute the average diurnal cortisol level and the hourly decline in diurnal cortisol using linear regression, which then entered the third analysis.

### Glucocorticoid pathway gene expression in T cells

In the second study project, we measured the gene expression of four key components of GC signaling, FK506-binding protein 4 (FKBP4) and FKBP5, GC-induced leucine zipper (GILZ), and the GC receptor (GR), separately for CD4^+^ and CD8^+^ T cells. Acting as a co-chaperon, FKBP5 modifies the activity of the GR, the essential receptor for immunomodulation, and thereby modulates the stress response in the immune system (39). FKBP5 is replaced by FKBP4 upon GC binding to the GR, which initiates nuclear translocation and subsequent transcriptional activity. Lastly, GILZ is transcriptionally activated by GR and initiates crucial anti-inflammatory GC pathways, especially in T cells (40). Using a real-time PCR system, complementary DNA was amplified in order to measure the expression of these four markers in the T cell subpopulations. The gene expression was then normalized to that of housekeeping genes by computing the delta cycle threshold (ΔCT; defined as gene of interest geometric mean CT – housekeeping genes geometric mean CT). GC sensitivity markers were available from 37 PwMS (see Supplement for more information, including specifics on the isolation of peripheral blood mononuclear cells, the sorting of CD4^+^ and CD8^+^ T cells, RNA isolation, cDNA synthesis, and real-time PCR).

### Statistical analysis

#### Analysis 1: BMI and MS severity

We employed robust linear models to regress four criteria of interest (CI; i.e., the annualized relapse rate, clinical disability, whole-brain T2-weighted lesion load, and GM fraction) on BMI in four independent analyses. Sex, age, study project, severity of depressive symptoms, disease duration, the presence of a progressive MS type, and disease modifying treatment were modeled as covariates of no interest (CNI). Additionally, from the four CI, the three CI not tested in the respective CI-specific analysis were also modeled as CNI. Significance was evaluated with permutation testing in undirected tests. Here, and in all other permutation analyses 10,000 permutations were conducted. An FWE-corrected significance threshold of α_FWE_ = 0.05 corresponding to 0.05 / 4 = 0.0125 on a single test level was applied in undirected tests. We report Cohen’s effect size measure f^2^ (f^2^ ≥ 0.02 weak, f^2^ ≥ 0.15 medium, and f^2^ ≥ 0.25 strong effect; [41]).

#### Analysis 2: BMI and functional connectivity of stress-reactive brain regions

For testing associations between BMI and FC of stress-reactive brain regions, we determined brain regions sensitive to stress on the group level in a first step and then related the stress and rest stage FC between stress-reactive regions to BMI across patients in a second step. Specifically, using the regression coefficients described in “Regional neural stress reactivity and functional connectivity within patients” as dependent variables, we employed robust linear regression to test for differences in the regions’ activity during stress vs. rest across patients. CNI were all variables evaluated in the first analysis. In addition, cognitive task load (i.e., the average inter-trial period in the final eight minutes of the stress stage) and time-to-feedback (i.e., the duration of the “Evaluation” substage) were modeled as stress-task-specific CNI. Modeling cognitive task load is crucial to control for putative effects of task demands and mental arithmetic performance. Statistical significance was evaluated by permutation testing. A family-wise-error (FWE) corrected significance threshold of α_FWE_ = 0.05 was applied in undirected tests corresponding to 0.05 divided by 119 (number of brain regions included in the analysis) or 4.2 · 10^−4^ respectively on a single test level. An analysis of perceived psychological stress reactivity (i.e., self-report data) and of peripheral autonomic stress reactivity (i.e., heart rate) can be found in the Supplement (Supplementary analysis 1). After stress-reactive regions were identified, we regressed the FC of each pair of stress-sensitive regions on BMI using robust linear regression, separately for the rest and the stress stage FC. CNI were identical to those used for the identification of stress reactivity. Permutation testing was employed for undirected pair-of-region-wise tests of significance. Given that 17 [17 - 1] / 2 = 136 tests were conducted per condition, we applied an α_FWE_ = 0.05 equal to 0.05 / (17 [17 - 1] / 2) = 3.68 · 10^−4^ on a single test level. Notably, we conducted a supplementary analysis which regressed the neural stress reactivity parameters (i.e., the abovementioned coefficients) of stress reactive regions on participants’ BMI (Supplementary analysis 2) to help evaluate the relative suitability of FC for BMI prediction performed in analysis 2.

#### Analysis 3: BMI and glucocorticoid functioning

Here, we tested associations between BMI and (i) the average diurnal cortisol level and (ii) the hourly decline in the diurnal cortisol level (α_FWE_ = 0.05 equal to 0.05 / 2 on a single test level). Moreover, we tested potential links between BMI and the expression of FKBP4 and FKBP5, GILZ and the GR in CD4^+^ and CD8^+^ T cells separately. Correspondingly, we applied α_FWE_ = 0.05 (equal to 0.05 / 8 on a single test level) to test for significance and used the same CNI as in the second analysis except for the two stress-task specific CNI (cognitive task load and time-to-feedback).

### Data availability

Anatomical MRI images cannot be shared due to privacy regulations for clinical data use. Other data used in this study is available on reasonable request depending on the ethics committee’s approval and under a formal Data Sharing Agreement.

## Results

### Clinical and demographic participant characteristics

Six PwMS were obese (kg/m^2^ ≥ 30), 19 over- (25 ≥ kg/m^2^ > 30), 28 normal- (18.5 ≥ kg/m^2^ > 25), and four underweight (kg/m^2^ < 18.5). Five patients were treated with teriflunomide, ten with dimethyl fumarate, nine with fingolimod, eight with glatiramer acetate, and ten with β-interferons. See Tab. 1 for further information.

**Table 1.** Demographic and clinical patient characteristics. Abbreviations: ARR, annualized relapse rate; BDI, Beck Depression Inventory; DD, disease duration since first signs; EDSS, Expanded Disability Status Scale; f., female; GMF, whole-brain grey matter fraction; m., male; PROG, presence of progressive MS; pts., points; rel., relapses; T2LL, whole-brain T2-weighted lesion load; yr., year; yrs., years.

| **Sex**<br>(f./m.) | **Age**<br>(yrs.) | **BMI**<br>(m/kg<sup>2</sup>) | **BDI**<br>(pts.) | **EDSS**<br>(pts.) | **DD**<br>(yrs.) | **ARR**<br>(rel./yr) | **PROG**<br>(y/n) | **T2LL**<br>(cm<sup>3</sup>) | **GMF** |
| --- | --- | --- | --- | --- | --- | --- | --- | --- | --- |
| # | MN<br>SD | MN<br>SD | MN<br>SD | MD<br>RG | MN<br>SD | MN<br>SD | # | MN<br>SD | MN<br>SD |
| 37 | 46.4 | 24.7 | 8.2 | 2.5 | 12.8 | 0.62 | 7 | 13.9 | 0.38 |
| 20 | 10.6 | 4.3 | 7.3 | 0 - 6 | 8.8 | 0.44 | 50 | 19.9 | 0.05 |

### Analysis 1: BMI and multiple sclerosis severity

For the four severity parameters tested, the association between BMI and relapse rate was significant on an FWE-corrected level of α_FWE_ = 0.05 (equal to 0.05 / 4 = 0.0125 on the single test level). In particular, we obtained t = 3.23, p = 0.003, and f^2^ = 0.22 for this measure. We did not find significant links to any of the other parameters investigated (EDSS: t = -0.88, p = 0.378, f^2^ = 0.01; lesion load: t = 0.26, p = 0.792, f^2^ = -0.00; GM fraction: t = 0.24, p = 0.812, f^2^ = -0.01; see Fig. 2).

**Figure 2.**
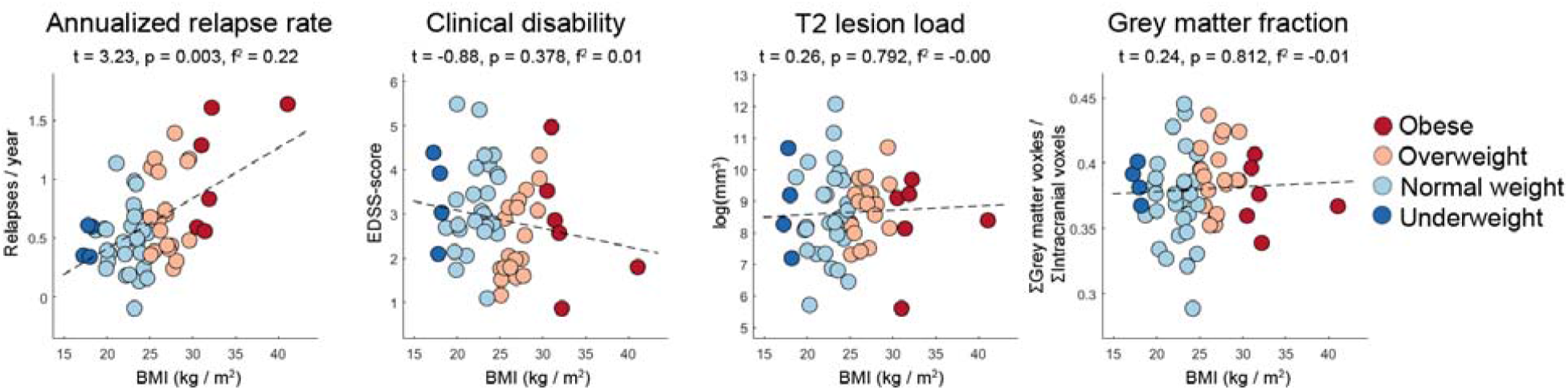
Associations between BMI and multiple sclerosis severity. The scatter graphs illustrate the associations found in analysis 1 for BMI and the four MS severity markers. To facilitate a more comfortable interpretation of body mass in these and all other scatter graphs, the dot colors highlight BMI-based body mass classes, with red dots corresponding to obese PwMS (kg/m^2^ ≥ 30), pink to overweight PwMS (25 ≥ kg/m^2^ > 30), light blue to normal weight PwMS (18.5 ≥ kg/m^2^ > 25) and darker blue dots to underweight patients (kg/m^2^ < 18.5). Noteworthy, however, these classes are only depicted for illustrative purposes, the regression models (across all analyses in the main text and the supplement) evaluated continuous BMI scores.

### Analysis 2: BMI and Functional Connectivity

Seventeen widely distributed brain regions showed significant stress–rest activity differences and were thus considered stress-reactive (Fig. 3a). BMI was significantly linked to FC between right anterior insula and right supramarginal gyrus during the rest stage (t = 4.02, p = 2.5 · 10^−4^, f^2^ = 0.51) according to α_FWE_ = 0.05 (α_single test_: 0.05 / 136 = 3.68 · 10^−4^). For stress stage FC, we found a significant association between right cerebellum exterior and right superior parietal lobule (t = -3.67, p = 3.3 · 10^−4^, f^2^ = 0.30; see Fig. 3b – d for an illustration).

**Figure 3.**
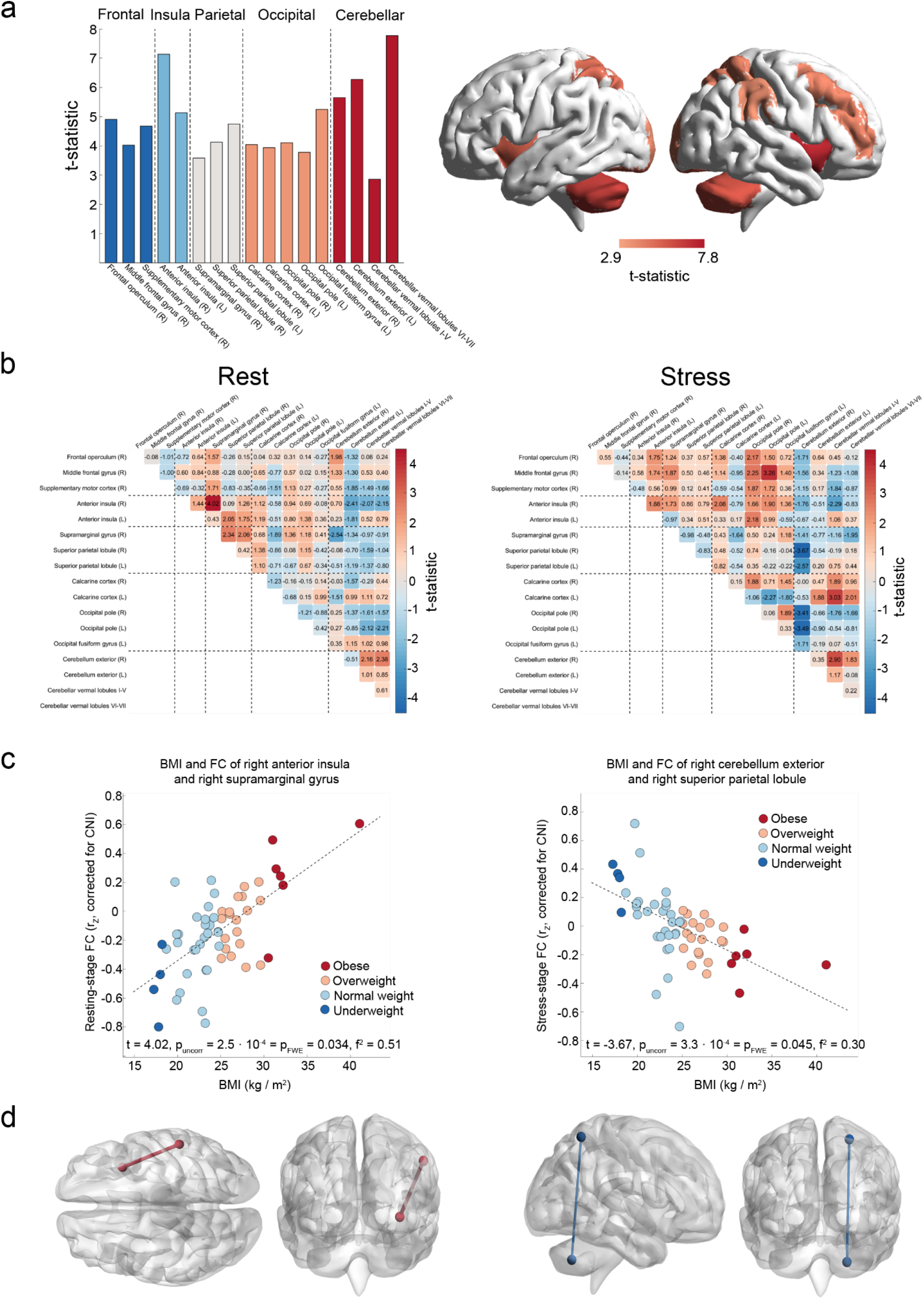
BMI and functional connectivity of stress-reactive brain regions. 3a depicts the 17 stress-reactive brain areas (i.e., with significant activity differences between the rest and stress stage) identified. The bar graph on the left depicts the t-statistics obtained for the stress – rest contrast for significant regions sorted by their coarse anatomical location. The two graphs in the middle/right of 3a correspond to surface renderings of regional stress reactivity. The heatmaps in 3b depict associations between BMI and FC of pairs of stress-sensitive regions separately for FC during the rest and the stress stage. The left scatter graph in 3c illustrates the association between the pair of regions with significant rest stage BMI associations according to a threshold corrected for multiple comparison (α_FWE_ = 0.05; right anterior insula to right supramarginal gyrus). The scatter graph on the right shows the same for the pair with a significant BMI to stress stage FC association (right cerebellum exterior to right superior parietal lobule). Finally, 3d depicts the median coordinates of illustrative connections between these regions.

### Analysis 3: BMI and glucocorticoid functioning

The association between BMI and the hourly decline in salivary cortisol barely failed to reach significance according to α_FWE_ = 0.05 (α_single test_: 0.05 / 2 = 0.025) as we computed t = - 2.28 (p = 0.031, f^2^ = 0.01) for this link. However, a significant link to BMI was obtained for the expression of the GC sensitivity markers FKBP4 (t = -2.96, p = 0.003, f^2^ = 0.13) and FKBP5 (t = 1.83, p = 0.003, f^2^ = 0.38) in CD8^+^ cells (Fig. 4) according to α_FWE_ = 0.05 (α_single test_: 0.05 / 8 = 0.00625). An explanation for p = 0.003 given t = 1.83 (which follows from the combined application of robust regression and permutation testing) can be found in the supplementary Discussion.

**Figure 4.**
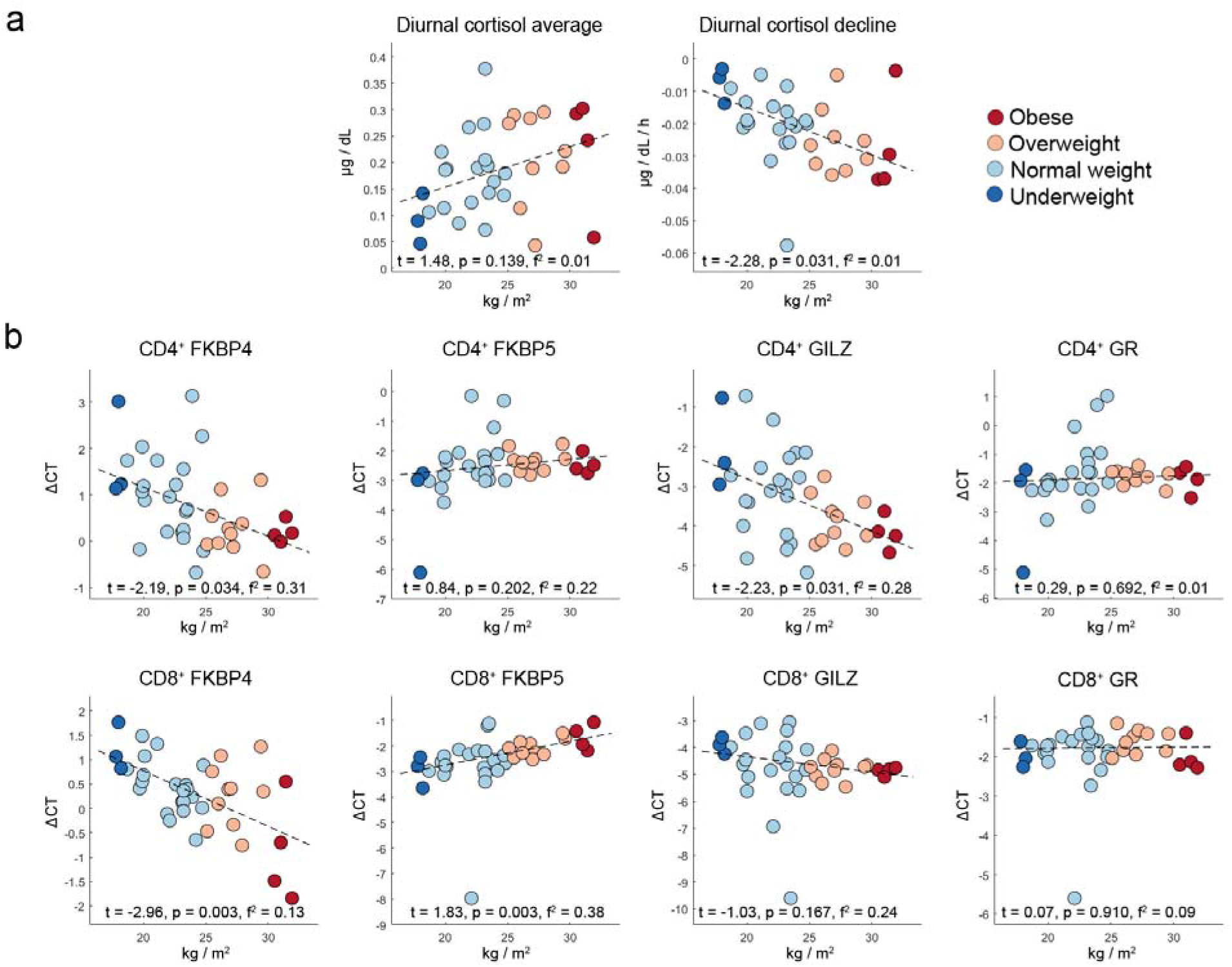
Associations between BMI and parameters of glucocorticoid functioning. The graphs in 4a show associations between BMI and the average diurnal salivary cortisol level on the left and between BMI and hourly decline in diurnal cortisol on the right. 4b depicts associations between BMI and the gene expression of four GC signaling-related markers in CD4^+^ and CD8^+^ T cells. Abbreviations: ΔCT: delta cycle threshold gene expression values; FKBP4, FK506-binding protein 4; FKBP5, FK506-binding protein 5, GILZ, glucocorticoid-induced leucine zipper; GR, glucocorticoid receptor.

## Discussion

Obesity is associated with increased disease severity in MS but important neurobehavioral obesity mechanisms including psychobiological stress have never been investigated. Consequently, we employed an fMRI stress task to study whether neural stress processing is related to body mass in 57 underweight to obese PwMS. Additionally, we tested whether body mass is related to salivary cortisol and GC sensitivity of T cells in patient subsamples. We found that BMI was linked to FC of anterior insula and supramarginal gyrus during rest and to FC between cerebellum exterior and superior parietal lobule during stress. Further, it was linked to gene expression of FKBP4 and FKBP5 on CD4^+^ and CD8^+^ T cells.

We conducted three main analyses: Analysis 1 served as proof-of-concept analysis testing whether BMI is linked to MS severity. Among the four severity measures tested (annualized relapse rate, clinical disability, T2-weighted lesion load, and GM fraction), a significant association was found for relapse rate. This is consistent with (2) who also tested relapse rate, EDSS and MRI parameters but only found an association for BMI and relapse rate. The authors provide the chronic low-grade grade level of inflammation induced by obesity and its impact on endothelial dysfunction as a potential explanation for this link.

Analysis 2 tested associations between BMI and FC of stress-sensitive regions. The first part of the analysis, which evaluated regional stress-reactivity, identified 17 regions (Fig. 3a) typically involved in stress processing (e.g., compare [22, 42]). Given that Supplementary analysis 1 showed that our task also induced a marked psychological and peripheral stress response, this underlines its ability to induce and measure psychological stress. The second part showed that, among these 17 areas, BMI was linked to FC of right anterior insula and right supramarginal gyrus during rest. This association is highly compatible with findings reported in the literature. In particular, insula is a key region of the incentive salience system (7) consistently showing enhanced activity to context stimuli of food (19). Insula and supramarginal gyrus are furthermore both parts of the so-called salience resting-state network (21, 43), whose function is to identify the most relevant among present stimuli for guiding subsequent behavior (43). Consistently, (21) reported increased activity of this network for persons with obesity. Thus, one can assume that the observed link reflects a positive relation between BMI and incentive salience of food stimuli.

For the stress stage, a significant negative relation between BMI and FC of right superior parietal lobule and cerebellum exterior was found. Van der Laan et al. (44) showed, using a food-choice fMRI task, that superior parietal cortex is a region involved in cognitive control as self-control (i.e., choice of low-caloric and healthy but not tasty foods) is accompanied by right superior parietal gyrus activity. Cerebellum is viewed as region much involved in the generation of stressful and affectively negative states (42), which is underlined by its distinct anatomical connections to other stress regions and its high density of cerebellar stress hormone receptors (42). Thus, considering that stress reduces the influence of control regions over regions favoring immediate reward within the goal-directed system and promotes unhealthy choices by reducing the regions’ FC (20), one can inversely assume that if a stress-state generating region adapts its activity to that of a control region (i.e., if the regions have a high FC), behavioral self-control increases and BMI decreases. Consequently, the observed negative relation presumably indicates the effects of self-control on BMI.

Notably, Supplementary analysis 2 investigated associations between BMI and activity of stress-reactive brain regions to help evaluating the relative suitability of FC for BMI prediction performed in analysis 2. Contrary to FC-based analysis 2, this supplementary analysis failed to reveal significant associations.

Analysis 3 evaluated relations between BMI and the hourly decline in and the average diurnal salivary cortisol as well as the expression of genes specific for CD4^+^ and CD8^+^ T cell GC sensitivity pathways. Although the analysis of both salivary cortisol parameters failed to reveal significant results on an FWE-corrected level, associations between BMI and FKBP4 and FKBP5 on CD8^+^ T cells were significant. This finding is consistent with results showing altered GC sensitivity in overweight and obese persons without MS by (14, 16) and underlines the importance of GC functioning for body weight and obesity.

An aspect worth discussing is the use of ASL fMRI for analyzing FC as BOLD fMRI is still the most widely used FC technique (28). However, two factors indicate that ASL is highly suitable for studying FC. First, the number of clinical ASL FC studies is rising in the last years (e.g., 45 - 48). When additionally considering that six ASL FC studies were already summarized in a review (28), this shows that the technique is used in a substantial body of research. Second, studies evaluating signal detection properties of ASL FC found that it reliably identifies established resting-state networks such as the salience and default-mode networks (49). Maybe more important, however, is the fact that ASL fMRI is much less affected by slow signal artifacts than BOLD fMRI (22). Thus, if a slow signal is observed, it can be retained because it can be attributed to the underlying experimental condition – in this case stress - which is a slow phenomenon (22). In BOLD fMRI, however, slow signal variations can either be induced by noise or by the experimental condition. Under these circumstances, removing slow signal variations is necessary but might remove the artifact and the experimentally induced signal (co-) variation as well.

Further, while this study’s sample size was not defined based on a sample size calculation, the present work evaluates a large number of samples compared to similar MS fMRI studies (i.e., according to a review [50], it evaluates a larger number of patients than 88% of all MS fMRI studies employing cognitive tasks).

A limitation of the study is that we did not standardize eating behavior on the day prior to the MRI scan. Moreover, although we modeled numerous potential nuisance factors in our analyses, smoking was not included. This may be considered another limitation due to interaction effects of smoking and BMI on MS disease parameters found in (2).

Taken together, our study demonstrates that stress-related processes in different biological systems are related to BMI in MS, which is in turn positively linked to relapse rate. Thus, in line with other studies (e.g., 25, 26), the present study underlines the importance of stress for MS severity and may in part elucidate underlying mechanisms.

## Author responsibility statement

Martin Weygandt takes full responsibility for the data, the analyses and interpretation, and conduct of the research; Martin Weygandt has full access to all data; Martin Weygandt has the right to publish any and all data, separate and apart from any sponsor.

### Statement on reporting guideline provided

The STROBE reporting guideline is provided with this submission.

### Statement on IRB or regional review board approval

The Methods section includes a statement that an IRB or regional review board has approved the use of humans for this study.

### Statement on permission obtained with regard to personal communication

The article does not include/cite personal communications.

### Statement on authors’ agreement with regard to conditions noted on the Authorship Agreement Form

All authors have agreed to conditions noted on the Authorship Agreement Form.

### Statement on participant’s consent forms

Martin Weygandt has received consent forms from any participant in the presented study and has them on file in case they are requested by the editor.

### Statement on participant’s consent forms allowing publication of figures or videos enabling participant recognition

No such figures or videos are included in this work.

