## Supplemental Material for "Neural stress processing, glucocorticoid functioning, and body mass in lean to obese persons with multiple sclerosis"

**SUPPLEMENTARY MATERIAL**

Short title: Obesity in multiple sclerosis

Keywords: Multiple sclerosis, obesity, psychological stress, functional connectivity, glucocorticoid functioning.

*These authors contributed equally

**Materials and methods**

**Heart rate computation**

We used the standard pulse oximeter of the Physiological Monitoring Unit included with the MRI scanner to determine the heart rate during rest and stress fMRI measurements. The photoplethysmography detector was mounted to the participants' toes as the experimental paradigm required them to manually operate button boxes. The quality of heart rate signals obtained from participants’ toes is comparable to signals obtained from their fingertips (Hinkelbein et al. 2005). We calculated a heart rate parameter for each participant and each of the two conditions. In particular, in the first study project (e.g., Weygandt et al., 2016), we removed pulse oximeter raw signal artifacts that would have resulted in a heart rate acceleration of ≥ 133% or a deceleration of ≤ 75% in a first step. The remaining data from the (final) 8 min of both conditions were then utilized to compute the average heart rate for a given participant and condition as indicator of heart rate of a given condition and participant in a second. In the second study project (Brasanac et al., 2022), we first computed a Fourier analysis to calculate the amplitude spectrum of heart rate time series based on the raw pulse signals of the (final) 8 min of both conditions separately for each condition and participant. To limit the fit to the physiologically meaningful frequency spectrum, frequencies below 30 Hz and above 150 Hz were eliminated. Amplitudes in two frequency windows (47.6 to 48.4 Hz, and 95.6 to 96.4 Hz) were also eliminated due to a technical artifact produced in these frequency ranges by the pulse oximeter. In a second step, we fitted a unimodal gaussian to the calculated amplitude spectrum for both periods. We then used the frequency with the largest fitted amplitude as the indicator of heart rate of a given condition and participant.

**MRI acquisition**

All brain scans were performed with a 3 Tesla whole-body tomograph (Magnetom Trio, Siemens, Erlangen, Germany) using a standard head coil with twelve channels.

Anatomical scans

In the first study project, two anatomical MR sequences were acquired. In particular, a T1-weighted sagittal 3-D-magnetization prepared rapid gradient echo (MP-RAGE) sequence (176 slices; slice thickness 1.3 mm; in-plane voxel resolution 1.5 · 1.5 mm^2^; TR = 1720ms; TE = 2.34ms; FA = 9°; FOV = 192 · 192 mm^2^; matrix size = 128 × 128; duration 1 min and 43 sec) and a sagittal T2-weighted (T2w) sequence to facilitate manual lesion mapping (176 slices; 1 mm isotropic voxels; TR = 5000 ms; TE = 502 ms; FA = 120°; FOV = 256 · 256 mm^2^; matrix size = 256 · 256; 5 min and 52 sec duration) was used. In the second project, we used a T1-weighted sagittal 3D MP-RAGE echo sequence (176 slices; 1 mm isotropic voxels; TR = 1900 ms; TE = 3.03 ms; FA = 9°; FOV = 256 · 256 mm^2^; matrix size = 256 × 256; 4 min 26 sec duration) as well as a sagittal T2-weighted FLAIR sequence (176 slices; 1 mm isotropic voxels; TR = 6000 ms; TE = 388 ms; TI = 2100 ms; FA = 120°; FOV = 256 · 256 mm^2^; matrix size = 256 · 256; 7 min 44 sec duration).

Functional scans

In both study projects, perfusion brain images were acquired using a pseudo-continuous ASL Echo-Planar Imaging (EPI) sequence (Wang et al., 2005). This sequence comprised 22 ascending transversal slices and covered the whole brain (slice thickness 5.75 mm [including 15% inter-slice gap]; in-plane voxel resolution 3 · 3 mm^2^; TR = 4000 ms; TE = 19ms; FA = 90°; FOV = 192 · 192 mm^2^; matrix size = 64 · 64; label duration 1.5 sec, post-label delay 1.2 s; phase-encoding direction anterior to posterior). Moreover, we acquired two spin-echo EPI reference volumes with opposing phase encoding directions (anterior to posterior, posterior to anterior) in advance to the rest and the stress ASL measurements with matching readout and geometry in order to facilitate a distortion correction of ASL images.

**MRI preprocessing**

Anatomical scans

First, experienced raters conducted a manual mapping of localized lesions based on the T2 or FLAIR scans. A neuroradiologist oversaw these procedures. Next, the T2 or FLAIR scan of a given participant was coregistered to this participants T1-weighted MP-RAGE scan. Afterwards, we determined voxel-wise tissue probability maps for grey matter (GM), white matter (WM) and cerebro-spinal fluid (CSF) by applying the combined spatial normalization and segmentation algorithm of SPM12 to T1-weighted MP-RAGE scans. Voxel locations included in the coregistered lesion masks were discarded in this combined normalization and segmentation procedure, which finally generated tissue probability maps for each of the three tissues. These (‘modulated’) tissue probability maps were adjusted for impacts of local deformations applied during spatial normalization and created in the anatomical standard space defined by the Montreal Neurological Institute (MNI). The transformation parameters determined in this procedure for mapping the T1-weighted images to the MNI space were also used to map the lesion masks coregistered to the space of the raw T1-weighted scans to the MNI space.

Afterwards, we used the modulated tissue probability maps to compute the whole-brain GM fraction for each patient. In particular, for each voxel, we identified the tissue with the largest modulated tissue probability in the given participant. We then assigned this voxel to the tissue identified except if its localization was within lesioned tissue as indicated by the MNI-space lesion mask. The GM tissue fraction was then calculated as the total number of GM voxels divided by the total number of intracranial voxels.

Similarly, group-masks for the three tissues (required in the analyses of functional MRI scans) were determined by computing the tissue with the largest modulated tissue probability across all participants and assigning it to this tissue. In order to account for partial voluming effects, we deleted coordinates from group masks that were directly located in lesioned tissue in at least one participant and also the six neighboring voxels (i.e., those six voxels that had a Euclidean distance of exactly one voxel to a lesion voxel of a given participant; cf. Weygandt et al., 2011).

**Quality assurance step for functional scans using the Framewise Displacement metric**

The framewise displacement (FWD) measure, a well-known technique proposed by Power et al. (2014) that assesses participants' fMRI head motion parameters (here determined during preprocessing of functional ASL scans), was used to perform an fMRI image data quality assurance step. Specifically, for each condition (i.e., rest and stress) and each of the 60 participants available (20 from Weygandt et al. (2016), 40 from Brasanac et al. (2022)), first one average motion-based picture quality marker ("FWD score") was calculated based on the head motion parameters determined during preprocessing of ASL scans. In the next step, we looked for outliers among the FWD scores across the pooled sample of 60 patients for each condition separately. In particular, an FWD score was deemed an outlier if more extreme than the first (third) quartile minus (plus) 1.5 times the inter-quartile range of the FWD scores computed for this condition. Only participants who had non-outlier FWD scores in both conditions passed the procedure. This was the case for all 20 participants taken from Weygandt et al. (2016), and for 37 from Brasanac et al. (2022).

**Regional neural stress reactivity**

In order to determine the regional neural stress reactivity, we first corrected the regional cerebral blood flow (rCBF) time series of each voxel, condition, and participant for head motion (Wang et al., 2012; Wang et al., 2008) and global WM and CSF signals (Arnemann et al., 2015; Wang et al., 2012). Head motion parameters were computed by ASLtbx, the average signal across all voxels contained in the group-specific WM and CSF masks were computed for each participant as global WM and CSF signals.

Next, we determined an rCBF timeseries for each condition, participant, and region included in the Neuromorphometrics brain atlas (http://Neuromorphometrics.com) averaged across timeseries of all voxels in that region if the given voxel was included in the GM group mask and contained non-zero rCBF. An atlas region was excluded from all fMRI analyses if not containing a single voxel in one or more participants meeting these criteria. From the 122 regions in the atlas, only three, i.e., left and right pallidum as well as cerebellar vermal lobules VIII-X, were not included. We excluded these regions as (i) the GM contrast in the pallidum is relatively weak in MP-RAGE scans (Droby et al., 2021), (ii) MS lesions occur frequently close to pallidum, (iii) the location of cerebellar vermal lobules VIII-X was close to the inferior boundary of the ASL measurement region (i.e., its Field of View), (iv) we excluded regions already if they did not fulfill the abovementioned criteria in a single participant.

To finally compute measures of regional stress reactivity, we concatenated both conditions’ regional time series in each participant and region and used linear regression to determine regional activity differences between the stress and the rest condition. The concatenated time series served as dependent variable and a boxcar regressor coding zeros for the 60 rCBF data points of the rest condition and ones for the 60 rCBF data points of the last 8 min of the stress condition served as predictor (e.g., Wang et al., 2012; Wang et al., 2008). This procedure was carried out for each person and region included in the analysis. The region-by-region regression coefficients obtained were entered into the second analysis as regional stress reactivity indicators.

**Gene expression in T cellular glucocorticoid pathway**

Isolation of Peripheral Blood Mononuclear Cells

A venous blood sample (about 70ml) was taken from participants at the morning of the clinical examination and processed within two hours. Particularly, in accordance with standard operating procedures (Hasselmann et al., 2018), peripheral blood mononuclear cells (PBMCs) were isolated with density gradient centrifugation. Specifically, blood was diluted first in phosphate-buffered saline (PBS) (1:1). Afterwards, 35ml of the diluted blood were layered carefully in a 50ml conical tube on top of 15ml of Biocoll medium (Biochrom, Berlin, Germany). The tubes were centrifuged at 870 x g (brakes off) for 30 min. After the mononuclear cell layer was harvested from the interphase, cells were washed twice in cold PBS for 10 min each. In order to prepare the PBMCs for cryopreservation, they were pelleted and then resuspended in RPMI-1640 (Gibco, ThermoFisher Scientific, Berlin, Germany), which was supplemented with 25% heat-inactivated fetal bovine serum (FBS; Biochrom, Berlin, Germany) and 10% dimethylsulfoxide (Applichem GmbH, Darmstadt, Germany). Next, cells were counted and then diluted to a concentration of 10 million cells per milliliter in 1.5ml tubes (Eppendorf, Hamburg, Germany), and placed in a freezing container (Sigma-Aldrich, St. Louis, USA) for overnight cooling at -80 °C. Frozen tubes were moved to a long-term liquid nitrogen storage tank (-196 °C) the following day where they remained until further examination.

Cell sorting, RNA isolation, cDNA synthesis, and real-time PCR

Magnetic-activated cell sorting (MACS), which divides cells based on surface antigens, was used to separate CD4^+^ and CD8^+^ T cell subsets. After thawing in a warm water bath (37 °C) for one to 2 min, tubes containing PBMCs were washed for 6 min at 250 × g in warmed RPMI-1640 medium containing 10% FBS. Following the manufacturer's recommendations, cell sorting was then carried out using CD4 and CD8 MicroBeads (Miltenyi Biotec, Bergisch Gladbach, Germany). 20µl of CD4 MicroBeads (Miltenyi Biotec, Bergisch Gladbach, Germany) and 80ml of MACS buffer (phosphate-buffered saline, 0.5% BSA, 2 mM EDTA) per 10 million cells were added, and the mixture was incubated for 15 min at 4°C in the dark. After washing with MACS buffer for 5 min at 350 × g (4 °C) cells were resuspended in 500µl MACS buffer. Next, MACS LS columns (Miltenyi Biotec, Bergisch Gladbach, Germany) were used to isolate CD4^+^ T cells. The CD4^-^ negative percentage was then used for sorting CD8^+^ T cells in the following phase, adhering to a similar protocol: CD8 MicroBeads (Miltenyi Biotec, Bergisch Gladbach, Germany) were used to label the cells. The cells were then incubated on the MACS MS columns, separated, and ultimately resuspended in 500µl of MACS buffer. Using flow cytometry (FACSCanto II, BD Biosciences, New Jersey, USA), cell fraction purity was checked. The purity coefficients were 91.67% + 6.74 SD (0.89 SEM) for CD4^+^ T cells and 95.15% + 5.73 SD (0.76 SEM) for CD8^+^ T cells. Following the recommendations of the manufacturer, total RNA was extracted from CD4^+^ and CD8^+^ T cell fractions with the Qiagen RNeasy Plus Mini Kit (Qiagen, Hilden, Germany). A NanoDrop spectrophotometer (NanoDrop 2000c, Thermo Fisher Scientific, Berlin, Germany) was used to evaluate their purity and concentration. Complementary DNA (cDNA) was transcribed from RNA in the next step in accordance with the manufacturer's instructions using Thermo Fisher Scientific's RevertAid H Minus First Strand cDNA Synthesis Kit and kept at -20 °C. TaqMan Gene Expression Assays (Thermo Fisher Scientific, Berlin, Germany) were employed to amplify cDNA with a StepOne real-time PCR System for GR (Hs00353740 m1), FKBP5 (Hs01561006 m1), FKBP4 (Hs00427038 g1), and GILZ (Hs00608272 m1). Triplicates were used to run real-time PCR experiments. Two housekeeping genes, TATA Box Binding Protein (TBP; Hs00427620 m1) and Importin 8 (IPO8; Hs00183533 m1), were used to standardize gene expression. These housekeeping genes were chosen as reliable gene references for gene expression analyses in human T cells (Ledderose et al., 2011). By subtracting the geometric mean of housekeeping genes from the mean CT values of a given gene of interest, delta cycle threshold (∆CT) values were computed as markers of gene expression within the glucocorticoid pathway. These ∆CT values were entered in the third analysis for evaluation.

**Statistical analysis**

Supplementary analysis 1: Psychological and peripheral stress responses

We used linear mixed models to test perceived stress (based on self-report data acquired in both rating stages included in the analyses and available for all 57 PwMS) and peripheral stress responses (i.e., differences in the heart rate between the rest stage and the final 8 min of the stress stage; available for 51 PwMS). The CNI were the same as in the second analysis. Again, we applied permutation testing for inference.

Supplementary Analysis 2: BMI and brain activity of stress-responsive regions

Here, we repeated the second analysis but instead of regressing the participants FC on their Body Mass Index (BMI), we modeled the regional stress response activity parameters (i.e., the coefficients described in section ‘Modeling regional neural stress reactivity and functional connectivity within patients’ of the main text) based on their BMI. CNI were the same as in the second analysis. Again, we applied a significance threshold corrected for multiple testing of α_FWE_ = 0.05. However, the equivalent of this FWE-corrected threshold on the single test level was 0.05/17 = 0.0029 as the number of tests equaled to the number of significant stress responsive regions (i.e., 17) and not the number of pairs of significant stress responsive regions (i.e., 136).

**Results**

**Supplementary analysis 1: Psychological and peripheral stress responses**

The task induced a pronounced psychological (t = 9.31, p < 10^-4^, f^2^ = 0.99) and peripheral (t = 6.05, p < 10^-4^, f^2^ = 0.71) stress response. Fig. S1 provides further details.

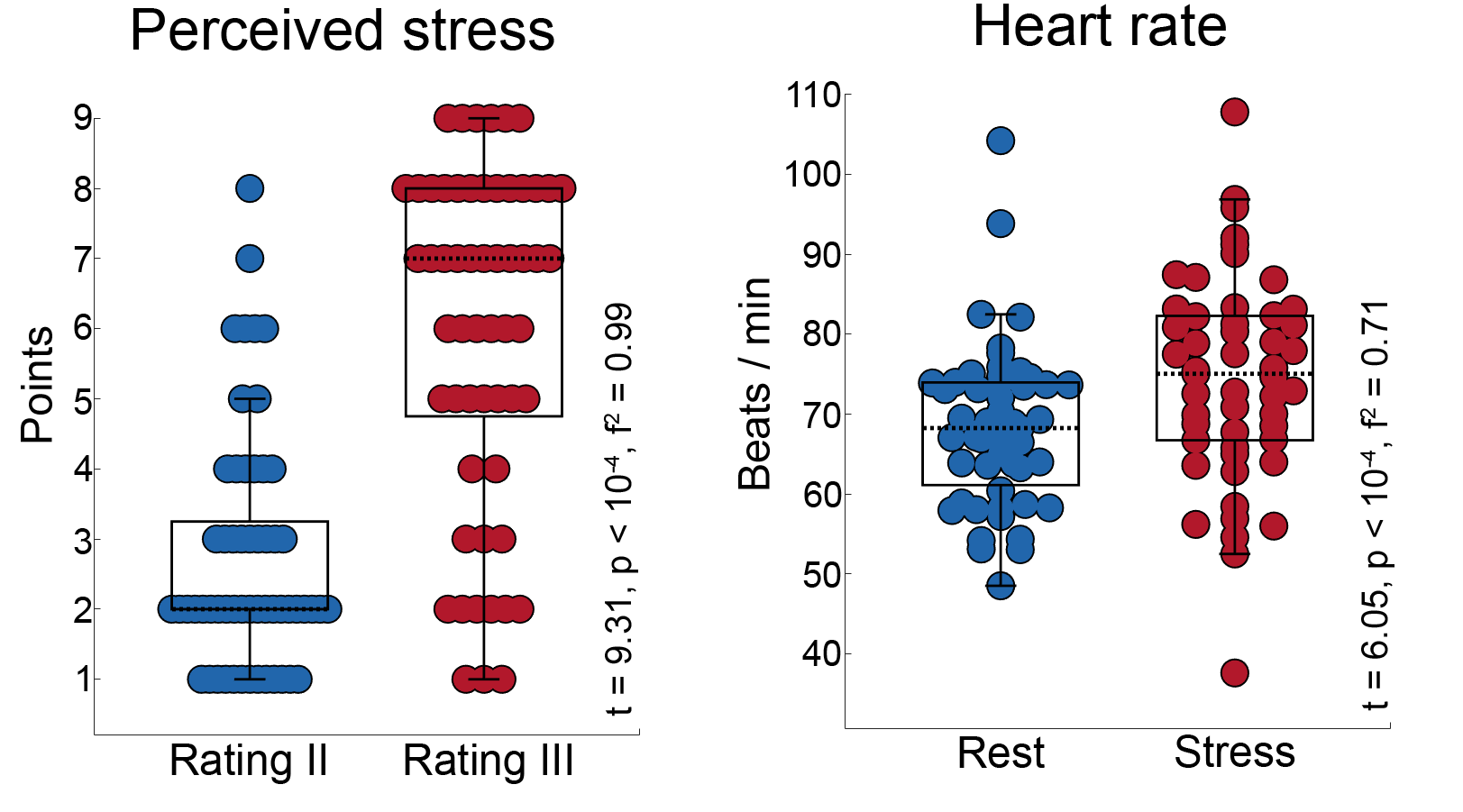

**Figure S1** depicts the stress response induced by the task in terms of psychological stress (ratings of perceived stress) on the left and in terms of heart rate accelerations on the right. The lower, center, and upper edges of the black boxes depict the first, second (i.e., median), and third quartile of the distribution of parameters.

**Supplementary analysis 2: BMI and brain activity of stress-responsive regions**

We found for none of the 17 regions a significant association between BMI and stress-response activity. Specifically, for the most extreme t-statistic obtained in this analysis (i.e., for the association between BMI and right anterior insula activity differences), the type I error rate was p = 0.01, which is much larger than the required p-value of 0.0029. Fig. S2 provides an illustration.

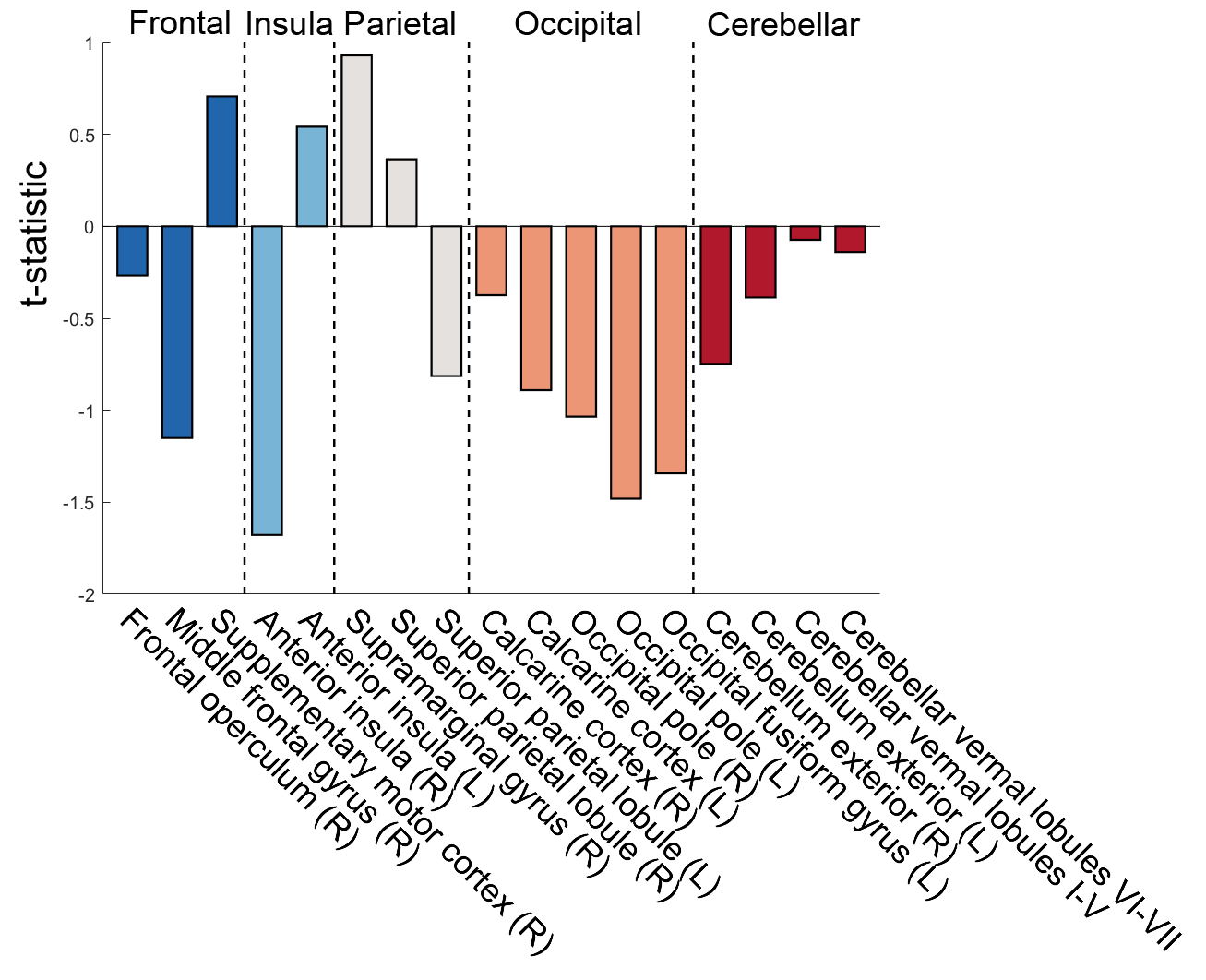

**Figure S2** illustrates the associations between the BMI and the regional stress response activity of stress-reactive regions.

**Discussion**

A strength of the study is the combination of robust regression and permutation testing, which is e.g., demonstrated by the analysis of BMI – CD8^+^ FKBP5 associations (Fig 4b). In this case, the t-statistic of 1.83 appears comparably small for a p-value of 0.003. This apparent mismatch is exactly an effect of combining the two methods. In particular, in regression, t-statistics are computed by dividing the coefficient of a predictor by its standard error. As robust regression is able to suppress the impact of outliers on the model, the estimated coefficient for BMI effects in the referred analysis is 0.094 for the depicted model including the outlier *and* for a model where this outlier is excluded. As it is impossible to suppress the impact of the outlier on the standard error (primarily governed by the data), though, the standard error is larger for the model with (0.051) than without the outlier (0.030). Consequently, the t-statistic for the model with outlier is t = 1.83 and, according to a parametric t-distribution, p = 0.08. The situation changes when the outlier is excluded: Under these conditions t = 3.15 and, again according to a parametric t-distribution, p = 0.004. Thus, under these conditions, the association between BMI and CD8^+^ FKBP5 is close even according to the parametric setting – but the parametric method is only able to detect this when no outliers are present – as parametric t-distribution are agnostic to their impact. Permutation testing, however, generates a t-distribution that adapts to the given data by permuting the predictor (here including an outlier) and thus reflects their impact. Due to these properties, the probability for obtaining the observed BMI effect when including the outlier determined by permutation testing is p = 0.003 and thus close to that of 0.004 determined by the parametric approach when the outlier is excluded.
